# Dynamics of abdominal symptoms during the start of a new therapy with Elexacaftor/Tezacaftor/Ivacaftor using the novel CFAbd-day2day questionnaire

**DOI:** 10.1101/2023.07.31.23293088

**Authors:** Jochen G. Mainz, Anton Barucha, Pu Huang, Lilith Bechinger, Franziska Duckstein, Louise Polte, Pauline Sadrieh, Lutz Nährlich, Olaf Eickmeier, Suzanne van Dullemen, Patience Eschenhagen, Carsten Schwarz, Stefan Lüth, Carlos Zagoya, Ute Graepler-Mainka

## Abstract

**Background:** Elexacaftor-tezacaftor-ivacaftor (ETI) is a novel highly effective CFTR modulator combination proven to improve lung function and body weight in people with Cystic Fibrosis (pwCF) carrying a F508del mutation. Recently, we revealed significant reductions of abdominal symptoms (AS) in German, British and Irish pwCF after 24-26 weeks of ETI using the CFAbd-Score, the first PROM specifically developed and validated for pwCF following FDA guidelines. Notably, many pwCF reported marked changes in their AS during the first days of the new treatment. To capture these immediate effects, we developed the CFAbd-day2day, a CF-specific GI-diary, following FDA and COSMIN guidelines.

**Aims:** To prospectively capture immediate dynamics of AS using the CFAbd-day2day 14 days before and 14-28 days after ETI initiation. In addition, we aim here to provide validation steps of the novel PROM concerning sensitivity to changes.

**Methods:** To develop the CFAbd-day2day, focus groups (community voice=pwCF and their proxies and CF specialists from different fields) were repeatedly consulted. Before and under the new ETI therapy pwCF prospectively scored AS on a daily basis with the CFAbd-day2day.

**Results:** Altogether, n=45 pwCF attended in 5 CF centers prospectively completed the CFAbd-day2day before (mean±sd: −14±7 days) and after (mean±sd: 28±23 days) ETI initiation. Whereas cumulative scores significantly decreased during the 3-4-week time frame after ETI initiation, compared to the two weeks prior to therapy, many patients who revealed a relatively stable level of AS before ETI reported changes during the first days of treatment with the highly effective CFTR modulators. Items like pain and flatulence increased in up to 21% of patients during the first 14 days of therapy but they improved during days 15-27.

**Conclusion:** The CFAbd-day2day, specifically developed and in process of validation to prospectively capture GI symptoms in pwCF, provides new substantial insights into the dynamics of AS in pwCF receiving a new treatment with ETI. The novel tool is also helpful to prospectively monitor patients with specific GI problems. International implementation and further validation steps of the diary are ongoing.

## 1 Introduction

For long, abdominal involvement in Cystic fibrosis (CF), the most common lethal inherited disease of the Caucasian population, received little attention. Since availability of pancreatic enzyme supplementation therapy (PERT) in mostly (1) pancreatic insufficient patients, pulmonary infection and lung destruction became the major reason for premature death in about 90% of people with CF (pwCF)(2,3). The causative gene defect results in abnormal production and function of the cystic fibrosis transmembrane conductance regulator (CFTR) protein, crucially affecting both the respiratory and gastrointestinal systems. Apart from the upper and lower airways, the ATP-gated anion channel is highly expressed in the pancreas, the gut and the bile ducts, leading to a CF-specific pattern of gastrointestinal (GI) complaints (4,5). Besides exocrine pancreatic insufficiency, present from birth in about 85% of our pwCF, impaired intestinal passage by meconium ileus, Distal Intestinal Obstruction Syndrome (DIOS) and constipation are typical complications in CF (6–8). These are complemented by factors like intestinal dysbiosis caused by frequent antibiotic treatments, cough-associated reflux, and maldigestion resulting in malresorption of nutrients which then allow bacterial fermentation, gases and diarrhea as principal symptoms of untreated exocrine pancreatic insufficiency (9). Finally, also endocrine liberation of insulin is hampered by the destruction of pancreatic beta cells, leading to substantial proportions of CF-related diabetes in pwCF during adulthood (10).

Introduction of highly effective CFTR modulator therapies (HEMT) in pwCF carrying a rare gating mutation like G551D over a decade ago revealed that correction and potentiation of the defective and/or malfunctioning CFTR channel has marked effects on the health status of pwCF beyond pulmonary function (11–14). Patients substantially gained weight and thrived if therapy was introduced in childhood (15). At the same time, from personal experience with our patients (16), we learned that HEMTs had effects on abdominal symptoms (AS). However, the lack of validated CF-specific PROMs focusing on GI involvement impeded former rigorous research to adequately assess the apparently substantial changes during early HEMT with ivacaftor in pwCF carrying a gating mutation.

To fill this gap, we developed the CFAbd-Score in different steps following FDA guidelines for development of a PROM (17) including CF patients and their families (community voice), as well as professional CF specialists at several time points. Initially, the PROM had been named JenAbd-Score (18), and after condensing it from a 5-sided questionnaire to a one-sided PROM, which comprises 28 items grouped in 5 domains, it was renamed “CFAbd-Score”(10,19–23). Presently, it is available in 9 languages and implemented in more than 25 studies around the world (22–26).

Recently, we implemented the CFAbd-Score in an international study with 107 pwCF from Germany and the UK before and at a median of 24 weeks during therapy with Elexacaftor-Tezacaftor-Ivacaftor (ETI). This HEMT was found to substantially improve the total CFAbd-Score and its 5 domains, i.e. “pain”, “GERD”, “disorders of bowel movement”, “disorders of appetite” and “quality of life impairment”(22). Quite a similar improvement pattern was observed during ETI in a parallel study administering the CFAbd-Score to 103 Irish and British pwCF before, as well as after 1, 2, 6 and 12 months of ETI (23). In both studies, the CFAbd-Score was observed to have high sensitivity to the changes induced by the new therapy with ETI, which is considered a game changer in CF. A recent multicenter study in the United States also found a significant reduction of GI symptoms in 263 pwCF receiving ETI. However, unlike results obtained with the CF-specific CFAbd-Score (22,23), changes assessed with questionnaires evaluated and validated for other GI pathologies, but not for pwCF [Patient Assessment of Upper Gastrointestinal Disorders-Symptom (PAGI-SYM), Patient Assessment of Constipation-Symptom (PAC-SYM) and Patient Assessment of Constipation-Quality of Life (PAC-QOL)] did not reach the level of clinical significance (27).

Nevertheless, during early phases after HEMT introduction, patients reported experiencing many GI symptoms, which may not be adequately represented by a PROM that addresses GI symptoms retrospectively with a two-week recall period. Furthermore, after introduction of the highly effective CFTR modulator, some symptoms could arise more frequently and possibly markedly, but only for quite a short period of time, which may not be adequately recorded with a questionnaire focused on capturing the overall frequency of GI symptoms over the past 14 days.

Consequently, we developed and are currently validating a prospective PROM as a diary, the CFAbd-day2day© questionnaire based on the CFAbd-Score, following FDA and COnsensus-based Standards for the selection of health Measurement INstruments (COSMIN) guidelines (17,28). Analogous to the development of the CFAbd-Score, pwCF and their families as well as professional CF specialists (community voice) were included to optimize wording and structuring of the PROM at several time points. The resulting prospective CF-specific GI-symptom diary CFAbd-day2day© is suitable for recording closely AS daily over a 14–day time period.

As the selection and wording of questions included in the CFAbd-Score has been found to be highly sensitive for capturing and quantifying GI symptoms in pwCF receiving a new ETI-therapy (22,23), substantial changes concerned adaptation of questions in terms of wording that allows prospective questioning of complaints on a daily basis. Furthermore, leaving room for individual comments and observations not captured by the questionnaire, i.e. eating habits, changes in PERT, etc. appeared essential for the CFAbd-day2day.

The aim of this study was to prospectively assess changes in abdominal symptoms on a daily basis during a new highly effective CFTR-modulating ETI therapy in pwCF. Therefore, symptoms should be captured in a period of up to 14 days prior to commencing the new therapy, as well as during 14-28 days of the new therapy using the novel CFAbd-day2day© questionnaire.

## 2 Methods

### 2.1 Participants

PwCF attending five German CF care centers in Brandenburg an der Havel, Potsdam, Tübingen, Gießen and Frankfurt am Main prospectively completed the CFAbd-day2day© before and after ETI initiation. Further criteria for inclusion were: confirmed evidence of CF by two positive sweat tests and/or detection of two CF mutations, carrying at least one allele with F508del, as requirement for ETI therapy initiation. Exclusion criteria were: inability to comply with the study procedures or assessments, and age below 6 years of age. Eligible subjects were included independent of their severity of pulmonary destruction (FEV1pred), airway colonization with specific pathogens and comorbidities. History of concomitant GI manifestations as well as food allergy or intolerance were quoted. Data acquisition was performed using *pseudonymization* and after written consent from the parents/legal guardian or from the pwCF themselves if age was above 18 years.

### 2.2 Assessment of Symptoms - CFAbd-day2day©

Abdominal symptoms were recorded daily using the CFAbd-day2day© questionnaire. The CFAbd-day2day© is a diary version of the CFAbd-Score. Therefore, the CFAbd-day2day© also comprises 28 items grouped into 5 domains. However, in addition to the modified recall period, some questions have been adapted to prospectively focusing on each observational day. The questionnaire also includes an optional comments section for recording changes in dietary habits, enzyme and medication use, as well as menstrual symptoms. A major intrinsic advantage of the diary character is the ability to record not only symptom frequencies but also their daily intensity. Printed copies of the questionnaires were administered to the participants by the local CF care providers and/or research coordinators. Scoring and analyses of completed, pseudonymized questionnaires were centrally performed at the CF center in Brandenburg an der Havel, Germany.

### 2.3 Statistical analyses

Individual baseline values for each pwCF for each of the 28 items included in the CFAbd-day2day© questionnaire were obtained by computing medians of each item over the time frame prior to ETI therapy initiation for each pwCF. Dynamics of symptoms were assessed by computing daily proportions of subjects reporting either improvement or worsening with respect to the above-defined baseline values for each of the 28 symptoms assessed with the CFAbd-day2day© questionnaire. Furthermore, within-subject averaged absolute daily variation measures were computed for each subject and each questionnaire item to quantify the levels of variation over three time frames, i.e. prior to ETI therapy, two and four weeks after ETI therapy initiation. These measures were computed by averaging the absolute changes experienced by each patient between consecutive days in each item over the previously mentioned time frames (see Supplementary material). Additionally, the maximum deviation with respect to the median in each item was identified for each patient within each time frame. In order to find out at which of the three above-mentioned periods of time symptoms reported by the cohort were observed to change the most, medians of these measures of variability were compared using non-parametric Wilcoxon signed-rank tests. Effects of ETI therapy on cumulative CFAbd-day2day responses were assessed by mapping the frequency of events registered by each patient for each item in the CFAbd-day2day© with the following scale: not at all → 0, once → 1, 2-3 times → 2, 4-7 times → 3, on more than seven days → 4 days and daily → 5. After that, items were grouped into five different domains as defined for the two-week CFAbd-Score (19). Scores for each domain and a total score were calculated for both the pre-ETI time frame and the two-week time period involving the third and fourth weeks after ETI therapy introduction. Comparisons of domains and total scores were conducted using non-parametric Wilcoxon signed-rank tests. Previous to these tests, normality assumptions were tested using quantile-quantile plots as well as Shapiro-Wilk tests.

## 3 Results

A total of 45 pwCF (median age: 10, [6-55] years) attended in each of the five participating CF centers (Brandenburg an der Havel, Potsdam, Gießen, Tübingen and Frankfurt am Main) were included in the study, of whom 30 (66.7%) were female and 15 (33.3%) male (Table 1). Patients included completed the diary for a mean of (mean±sd) −14±7 days and 28±23 days prior to and after commencing ETI therapy, respectively.

**Table 1.** Demographics and clinical characteristics of the pwCF included in this study.

|  |  |  |
| --- | --- | --- |
| **Demographics** | total (n) | 45 |
|  | age (median, range) | 10 (6-55) years |
|  | age < 18 | 37 (82.2) n (%) |
|  | age ≥ 18 | 8 (17.8) |
|  | female | 30 (66.7) |
|  | male | 15 (33.3) |
| **Genotype** |  |  |
|  | F508del homozygous | 25 (55.5%) |
|  | F508del heterozygous | 19 (42.2%) |
|  | F508del/unknown | 1 (2.2%) |
| **Previous CFTR modulator therapy** |  |  |
|  | Yes | 20 (44.4%) |
|  | No | 25 (55.5%) |
|  | lumacaftor+ivacaftor | 16 (35.5%) |
|  | tezacaftor/ivacaftor+ ivacaftor | 2 (4.4%) |
|  | ivacaftor | 1 (2.2%) |
|  | unknown | 1 (2.2%) |
| **CF associated diseases/conditions** |  |  |
| exocrine pancreatic insufficiency |  | Yes: 42/45 (93.3%)<br>No: 3/45 (6.7%) |
| meconium ileus |  | Yes: 8/45 (17.8%)<br>No: 37/45 (82.2%) |
| Distal Intestinal Obstruction Syndrome (DIOS) |  | Yes: 0 (0%)<br>No: 45 (100%) |
| CF-Related Diabetes (CFRD) |  | Yes: 3/45 (6.7%)<br>No: 36/45 (80%)<br>Unknown: (13.3%) |
| CF-Related Liver Disease (CFLD) |  | Yes: 9/45 (20%)<br>Liver fibrosis: 4/45 (8.9%)<br>Secondary biliary cirrhosis: 1/45 (2%)<br>Hepatic steatosis grade 1: 1/45 (2%)<br>Unspecified: 3/45 (6.7%)<br>No: 30/45 (66.7%)<br>Unknown: 6/45 (13.3%) |
| **Report on food allergies or intolerances** |  | Yes: 8/45 (17.8%)<br>No: 37/45 (82.2%) |
| **Lung function (FEV1 %pred) (conducted in 41 patients\*)** |  | 88.3±17.1% |
| **Body Mass Index (BMI)** |  |  |
| patients ≥ 18 years of age, n=8 (17.8%) | BMI (mean±sd) | 23.9 ± 4.0 kg/m<sup>2</sup> |
| patients < 18 years of age, n=37 (82.2%) | BMI-for-age z-Scores (mean±sd) | -0.34 ± 1.0 |

\* Baseline FEV<sub>1</sub>%pred values from four pwCF were not available.

### 3.1 Dynamics of symptoms

There was a high between- and within-subject variability in the responses of patients, and the dynamics over the considered time periods followed a rather individual profile in all 28 symptoms assessed with the CFAbd-day2day© (Figure 1).

**Figure 1.**
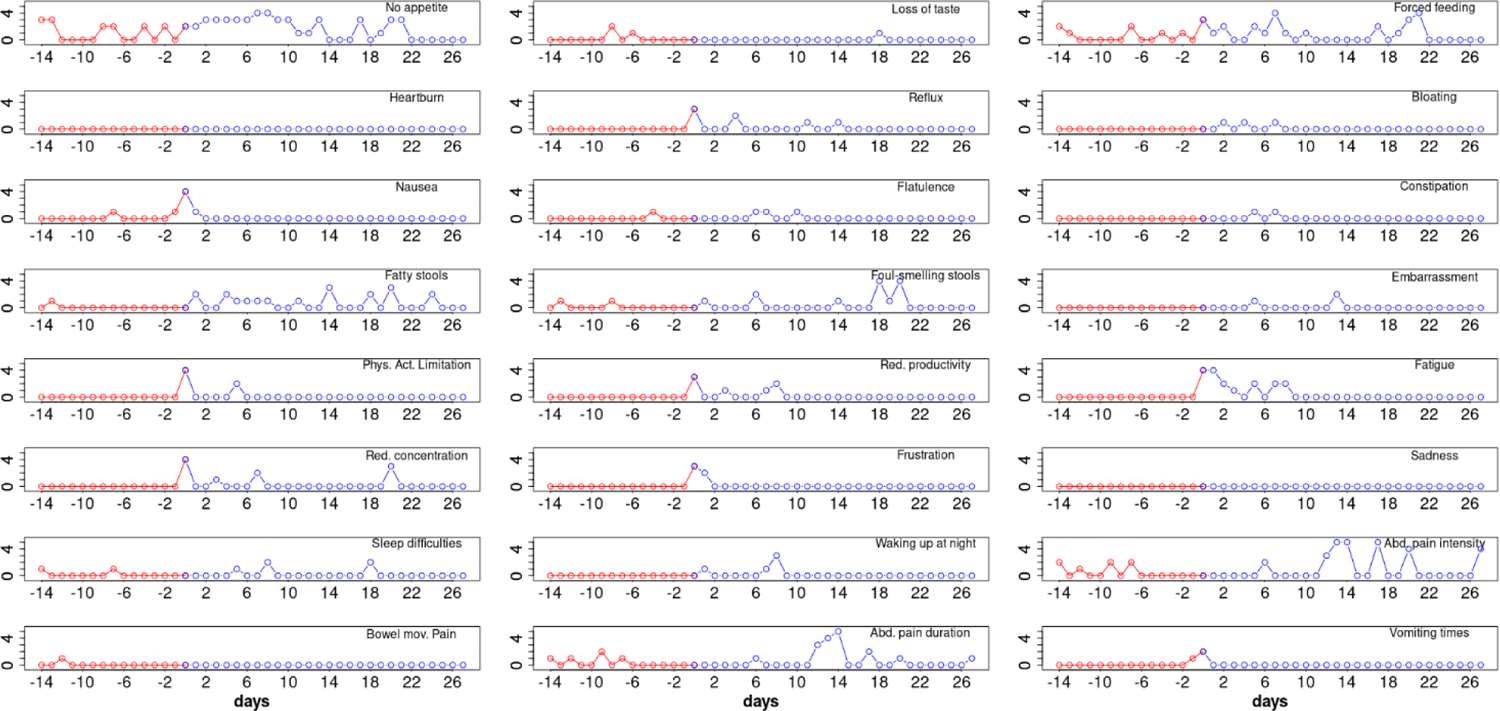
CFAbd-day2day^©^ protocol from a single CF patient (male, homozygous for F508del) revealing baseline symptoms before ETI (red) as well as evolution of the daily response during the first 27 days of therapy (blue) for 24 of the 28 items included in the CFAbd-day2day^©^ (y-axis represent score responses on a 0-5 Likert scale, where higher scores quote higher burden of symptoms).

Figure 2 shows proportions of patients reporting improvement with respect to the corresponding baseline value for each of the 28 items included in the CFAbd-day2day© over a maximum period of four weeks. As the question regarding “Pain intensity” depends on a positive answer for occurrence of “Pain”, these two questions were condensed into a single conditional question referred to as “Abdominal pain intensity” throughout the next sections of this article.

**Figure 2.**
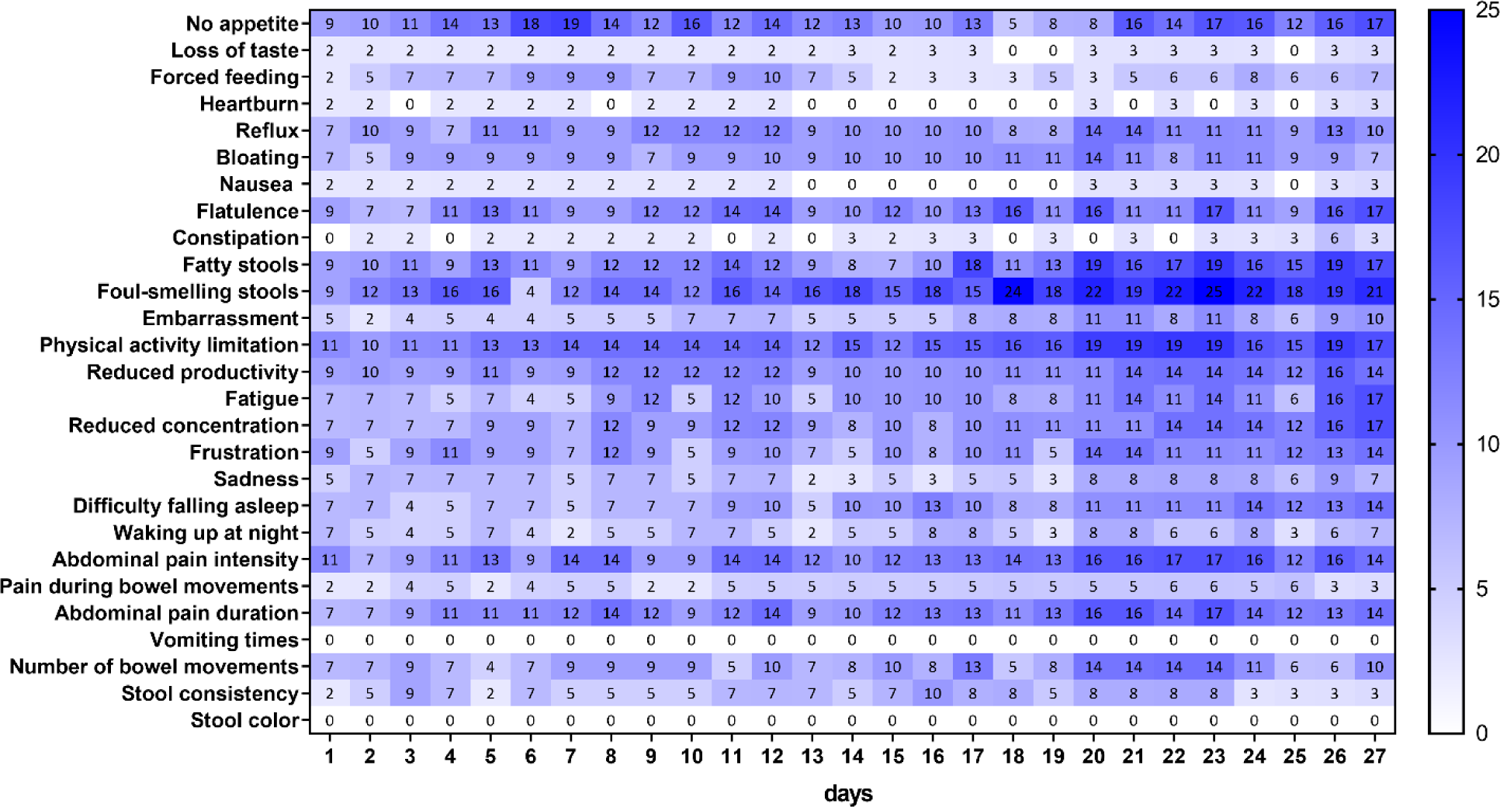
Proportion of pwCF reporting <u>improvement</u> for each item of the daily questionnaire with respect to their answers at baseline. Each percentage in the graph is relative to the total number of patients filling out the corresponding question on a specific day. Note that the questions regarding “Pain” and “Abdominal pain intensity” were condensed into a single conditional question.

In two items, i.e. “Vomiting times” and “Stool color”, no improvement with respect to baseline values was reported by the patients within this time frame. The highest proportion of patients (25%) reporting improvement was observed for the item “Foul-smelling stools” on day 23 after ETI therapy initiation. Other items for which the proportion of patients reporting improvement reaching almost the 20% level over this time frame were “No appetite” (19%), “Physical activity limitation” (19%) and “Fatty stools” (19%), with the former observed already on day seven and the other two from day 20 onward. On the other hand, symptoms for which the proportion of patients did not surpass the 10% level were “Loss of taste”, “Forced feeding”, “Heartburn”, “Nausea”, “Constipation”, “Sadness”, “Waking up at night” and “Pain during bowel movements”.

Regarding proportions of patients reporting worsening in symptoms within the first four weeks after start of ETI treatment (Figure 2), the highest was found for “Number of bowel movements”, for which 27% of patients on day 18 reported having a higher burden with respect to the time frame previous to ETI therapy. Second highest proportion was observed for “Flatulence” reaching the maximum (21%) on day 7. Interestingly, after ETI therapy initiation, one patient reported an increase in the number of “Vomiting times”, and two patients reported worsening in their “Stool color”. Other items with relatively low improvement, as reported by the patients over this time frame, were “Difficulty falling asleep”, “Waking up at night”, “Physical activity limitation” (in the three items max: 9%), “Loss of taste”, “Bloating” (in both max: 8%), “Reflux”, “Embarrassment”, “Sadness”, “Pain during bowel movements” (in all max: 7%), as well as “Heartburn” (max: 3%).

Table 2 shows averaged absolute daily changes in answers from patients reporting improvement or worsening in their GI symptoms (see Fig. 2 and 3) over the three considered periods of time, i.e. previous to ETI start, 1-14 and 15-27 days after ETI therapy initiation. According to this measure, variability levels for “Constipation” were significantly higher during the first two weeks after therapy initiation. On the other hand, the variability in “No appetite”, “Reflux”, “physical activity limitation”, “Abdominal pain intensity” and “Abdominal pain duration” was significantly lower during the third and fourth weeks after starting ETI therapy, compared to the time frame previous to commencing ETI. However, compared to the 1-14-day time frame, during the 15-27 days after ETI start, the variability in “No appetite”, “Reflux”, “Nausea”, “Reduced productivity”, “Fatigue”, “Reduced concentration”, “Difficulty falling asleep”, “Waking up at night”, “Abdominal pain intensity”, “Pain during bowel movements” and “Abdominal pain duration” was significantly higher during the first two weeks of ETI treatment.

**Figure 3.**
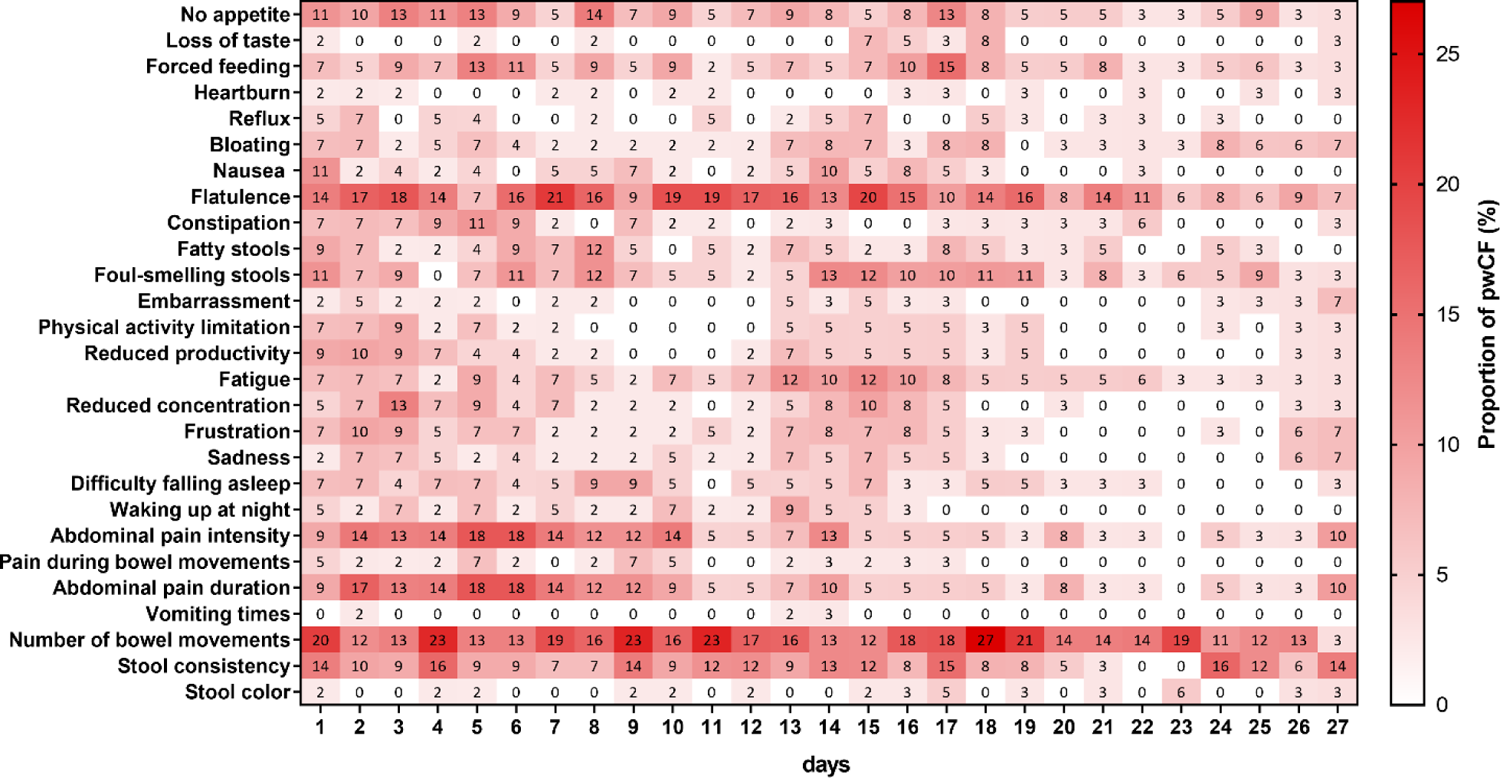
Proportion of pwCF reporting a <u>worsening</u> for each item of the daily questionnaire with respect their answers at baseline. Each percentage in the graph is relative to the total number of patients filling out the corresponding question on a specific day. Note that the questions regarding “Pain” and “Abdominal pain intensity” were condensed into a single conditional question.

**Table 2.** Averaged absolute between-day variability in answers from patients reporting improvement or worsening in their GI symptoms after ETI start (see Figs. 2 and 3). The post-ETI therapy period was split into two two-week time frames: 1-to-14 and 15-to-27 days after ETI therapy. Here, p1 indicates statistically significance for the comparisons of before and 1-to-14-day medians, p2 indicates statistically significance for the comparisons of before and 15-to-27-day medians, and p3 indicates statistically significance for the comparisons of 1-to-14-day and 15-to-27-day medians.

| Averaged absolute between day variability |  |  |  |  |  |  |  |
| --- | --- | --- | --- | --- | --- | --- | --- |
| Item name | n | T<sub>0</sub>:<br>14 days<br>before ETI<br>median, IQR | T<sub>1</sub>:<br>days 0-14<br>during ETI<br>median, IQR | T<sub>2</sub>:<br>days 15-28<br>during ETI<br>median, IQR | p<sub>1</sub><br>(T<sub>0</sub>-T<sub>1</sub>) | p<sub>2</sub><br>(T<sub>0</sub>-T<sub>2</sub>) | p<sub>3</sub><br>(T<sub>1</sub>-T<sub>2</sub>) |
| No appetite | 17 | 0.2 [0-0.7] | **0.4** [0.1-0.6] | **0.1** [0-0.3] | 0.82 | **0.02** | **0.02** |
| Loss of taste | 7 | 0 [0-0.2] | 0 [0-0.1] | 0.2 [0-0.2] | 1 | 0.83 | 0.35 |
| Forced feeding | 12 | 0.3 [0-0.4] | 0.4 [0.3-0.6] | 0.3 [0-0.3] | 0.4 | 0.33 | 0.07 |
| Heartburn | 7 | 0.3 [0.3-0.3] | 0 [0-0.4] | 0.3 [0.1-0.5] | 1 | 1 | 0.58 |
| Reflux | 13 | 0.2 [0.2-0.5] | **0.2** [0.1-0.3] | **0** [0-0.3] | 0.69 | **0.02** | 0.06 |
| Bloating | 14 | 0.2 [0-0.5] | 0.2 [0-0.4] | 0.1 [0-0.3] | 0.31 | 0.72 | 0.84 |
| Nausea | 13 | 0 [0-0.2] | **0.2** [0.1-0.3] | **0** [0-0.1] | 0.2 | 0.29 | **0.04** |
| Flatulence | 30 | 0.3 [0-0.7] | 0.4 [0.1-0.6] | 0.3 [0-0.5] | 0.89 | 0.49 | 0.59 |
| Constipation | 13 | **0** [0-0.1] | **0.2** [0.1-0.5] | 0 [0-0.1] | **0.04** | 0.59 | 0.23 |
| Fatty stools | 19 | 0.3 [0-0.6] | 0.2 [0.1-0.7] | 0.3 [0.2-0.6] | 1 | 0.66 | 1 |
| Foul-smelling stools | 21 | 0.2 [0-0.6] | 0.3 [0.1-0.6] | 0.4 [0.1-0.5] | 0.26 | 0.68 | 0.85 |
| Embarrassment | 10 | 0.4 [0.1-0.5] | 0.1 [0-0.4] | 0 [0-0.4] | 0.53 | 0.14 | 0.37 |
| Physical activity limitation | 16 | 0.3 [0.1-0.6] | 0.2 [0-0.3] | 0 [0-0.2] | 0.23 | **0.03** | 0.26 |
| Reduced productivity | 16 | 0 [0-0.2] | **0.2** [0.1-0.3] | **0** [0-0] | 0.1 | 0.07 | **0.01** |
| Fatigue | 16 | 0.3 [0-0.5] | **0.5** [0.2-0.8] | **0.2** [0-0.4] | 0.06 | 0.23 | **0.01** |
| Reduced concentration | 15 | 0.3 [0-0.4] | **0.3** [0.2-0.5] | **0.2** [0-0.3] | 0.41 | 0.2 | **0.03** |
| Frustration | 12 | 0.2 [0-0.7] | 0.2 [0.1-0.3] | 0.1 [0-0.3] | 0.73 | 0.08 | 0.11 |
| Sadness | 11 | 0.1 [0-0.4] | 0.2 [0.1-0.4] | 0.1 [0-0.2] | 0.69 | 0.27 | 0.39 |
| Difficulty falling asleep | 14 | 0.1 [0-0.4] | 0.2 [0.1-0.5] | 0 [0-0.2] | 0.36 | 0.14 | **0.004** |
| Waking up at night | 13 | 0 [0-0.3] | 0.3 [0.3-0.5] | 0 [0-0] | 0.23 | 0.09 | **0.01** |
| Abdominal pain intensity | 23 | 0.5 [0.1-0.9] | 0.4 [0.3-0.9] | 0.1 [0-0.3] | 0.59 | **0.01** | **0.01** |
| Pain during bowel movements | 7 | 0.1 [0-0.6] | **0.2** [0.1-0.5] | **0** [0-0] | 0.69 | 0.1 | **0.04** |
| Abdominal pain duration | 23 | 0.4 [0.1-0.6] | **0.4** [0.1-0.8] | **0.1** [0-0.3] | 0.81 | **0.003** | **0.003** |
| Vomiting times | 2 | 0 [0-0.1] | 0.4 [0.4-0.5] | 0 [0-0] | 0.5 | 1 | 0.5 |
| Number of bowel movements | 34 | 0.5 [0.1-0.8] | 0.5 [0.3-0.8] | 0.3 [0.2-0.7] | 0.85 | 0.32 | 0.09 |
| Stool consistency | 32 | 0.2 [0-0.7] | 0.3 [0-0.8] | 0.4 [0-0.7] | 0.94 | 0.41 | 0.5 |
| Stool color | 9 | 0 [0-0.1] | 0 [0-0.5] | 0.2 [0-1] | 0.28 | 0.14 | 0.37 |

P-values in bold represent statistically significant differences.

Comparing maximal absolute deviations from the median over the three time frames revealed that the maximum deviations in all items occurred within the first two-week period. This trend was statistically significant for items regarding “No appetite”, “Constipation”, “Fatigue” “Waking up at night” and “Abdominal pain duration” (Table 3). On the other hand, maxima in the “Foul-smelling stools” item observed within the first two-week period were significantly higher only when compared to the pre-ETI therapy time frame. For the five items: “Nausea”, “Reduced productivity”, “Difficulty falling asleep”, “Abdominal pain intensity” and “Pain during bowel movements” maxima occurring within the first two-week time frame were significantly higher compared to the those observed within the third and fourth weeks after ETI initiation.

**Table 3.** Comparison of maximal absolute deviations from the median observed in each patient reporting the changes observed in Figs. 2 and 3 within the time frames previous to ETI therapy, after 14 days and after 28 days of ETI therapy. The post-ETI therapy period was split into two two-week time frames. Here, p_1_ indicates statistically significance for the comparisons of before and 14-day medians, p_2_ indicates statistically significance for the comparisons of before and 28-day medians, and p_3_ indicates statistically significance for the comparisons of 14-day and 28-day medians.

| Maximal absolute deviations |  |  |  |  |  |  |  |
| --- | --- | --- | --- | --- | --- | --- | --- |
| Item name | N | T<sub>0</sub>:<br>14 days<br>before ETI<br>median, IQR | T<sub>1</sub>:<br>days 0-14<br>during ETI<br>median, IQR | T<sub>2</sub>:<br>days 15-28<br>during ETI<br>median, IQR | $p_1$<br>(T<sub>0</sub> -<br>T<sub>1</sub>) | $p_2$<br>(T<sub>0</sub> -<br>T<sub>2</sub>) | $p_3$<br>(T<sub>1</sub> -<br>T<sub>2</sub>) |
| No appetite | 17 | 1 [1-2] | 2 [1-3] | 1 [0-1] | **0.03** | 0.09 | **0.001** |
| Loss of taste | 7 | 0 [0-0.5] | 0 [0-1] | 1 [0.5-1.5] | 1 | 0.3 | 0.3 |
| Forced feeding | 12 | 1 [0-2] | 2 [1-3] | 1 [0-2] | 0.2 | 0.7 | 0.05 |
| Heartburn | 7 | 1 [0-2] | 1 [0-1] | 1 [1-1] | 1 | 1 | 0.8 |
| Reflux | 13 | 1 [1-2] | 1 [1-2] | 0 [0-1] | 0.8 | 0.07 | 0.1 |
| Bloating | 14 | 0.2 [0-1] | 1 [1-2] | 0.5 [0-2] | 0.4 | 1 | 0.3 |
| Nausea | 13 | 1 [0-3] | 2 [1-3] | 0 [0-0] | 0.3 | 0.2 | **0.03** |
| Flatulence | 30 | 1 [0-2] | 1 [1-2] | 1 [0-2] | 0.6 | 0.7 | 0.4 |
| Constipation | 13 | 0 [0-1] | 1 [1-2] | 0 [0-1] | **0.02** | 0.8 | **0.02** |
| Fatty stools | 19 | 1 [0-2] | 1 [0.5-2] | 1 [1-2] | 0.3 | 0.1 | 0.7 |
| Foul-smelling stools | 21 | 1 [0-1] | 2 [1-2] | 1 [1-2] | **0.02** | 0.2 | 0.6 |
| Embarrassment | 10 | 1 [0.1-2] | 1 [0-2] | 0.5 [0-2.5] | 0.5 | 0.9 | 0.9 |
| Physical activity limitation | 16 | 1 [0.4-2] | 1 [0-2] | 0 [0-1] | 0.6 | 0.2 | 0.08 |
| Reduced productivity | 16 | 1 [0-2] | 2 [1-2] | 0 [0-0.2] | 0.2 | 0.5 | **0.04** |
| Fatigue | 16 | 1 [0.7-1.2] | 2 [1-3] | 1 [0-1] | **0.03** | 0.5 | **0.01** |
| Reduced concentration | 15 | 1 [0-5-2.5] | 1.5 [1-2] | 1 [0-2.5] | 0.9 | 0.5 | 0.3 |
| Frustration | 12 | 1 [0-2] | 1 [1-2] | 1 [0-1] | 0.2 | 0.7 | 0.2 |
| Sadness | 11 | 1 [0-1.5] | 1 [1-2.5] | 0 [0-2.5] | 0.2 | 0.8 | 0.2 |
| Difficulty falling asleep | 14 | 1 [0-1] | 1.5 [1-2] | 0.5 [0-1] | 0.06 | 0.6 | **0.02** |
| Waking up at night | 13 | 0 [0-1] | 2 [1-2] | 0 [0-0] | **0.01** | 0.3 | **0.005** |
| Abdominal pain intensity | 23 | 2 [0.5-2] | 2 [1-3.5] | 0 [0-1.5] | 0.07 | 0.1 | **0.007** |
| Pain during bowel movements | 7 | 0.5 [0-2] | 2 [1-2] | 0 [0-0] | 0.09 | 0.4 | **0.03** |
| Abdominal pain duration | 23 | 1 [0.5-2] | 2 [1-3] | 0 [0-2] | **0.02** | 0.2 | **0.004** |
| Vomiting times | 2 | 1 [0.5-1.5] | 3.5 [3-4] | 0 [0-0] | 1 | 1 | 0.5 |
| Number of bowel movements | 34 | 2 [1-2] | 2 [1-2] | 1 [0.2-2] | 0.7 | 0.1 | 0.08 |
| Stool consistency | 32 | 1 [0-3] | 1.5 [0-3] | 1 [0-3] | 0.3 | 0.6 | 0.3 |
| Stool color | 9 | 0 [0-0] | 0 [0-3] | 3 [0-3] | 0.3 | 0.1 | 0.7 |

P-values in bold represent statistically significant differences.

**Table 4.** Changes in averaged CFAbd-day2day^©^ responses after ETI therapy introduction, comparing the cumulative burden of symptoms during the 14-day period before (T_0_) and the second 14-day period after initiation of the new therapy (T_2_: days 15-28).

|  |  | **T<sub>0</sub>:**<br>**14 days before ETI**<br>**median, IQR** | **T<sub>2</sub>:**<br>**days 15-28 during ETI**<br>**median, IQR** | **P** |
| --- | --- | --- | --- | --- |
| **Total cumulative CFAbd-day2day<sup>®</sup> scores** |  | 8.9 [2.8-15.7] | 4.7 [1.3-10] | **0.00001** |
| **Five Domains**<br>**of the CFAbd-Score** | **Pain** | 6.7 [0-20] | 0.0 [0-6.7] | **0.0003** |
|  | **GERD** | 0.0 [0-26.7] | 0.0 [0-6.7] | **0.01** |
|  | **Disorders of bowel movement** | 10.0 [2.5-22.5] | 7.5 [0-15] | **0.02** |
|  | **Disorders of appetite** | 4.0 [0-28] | 0.0 [0-20] | 0.1 |
|  | **Quality of life impairment** | 0.0 [0-5] | 0.0 [0-0] | **0.01** |

P-values in bold represent statistically significant changes between cumulative scores before and after ETI therapy.

### 3.2 Response to ETI therapy introduction

Effects of ETI therapy assessed with averaged CFAbd-day2day© responses reveals a highly significant decrease in the median total score (p=0.00001). Changes in four domains, i.e. “pain”, “GERD”, “disorders of bowel movement” and “quality of life impairment” were statistically significant (p=0.0003, 0.01, 0.02 and 0.01, respectively), although medians for “GERD” and “quality of life impairment” resulted equal for the two time frames.

## 4 Discussion

At this historic moment when highly effective CFTR modulators like ETI are changing the face of CF, it is essential to thoroughly identify the spectrum of effects of novel medications (13,29). At the same time, it may be a historic opportunity to capture the burden of symptoms in pwCF who, at baseline, are still naïve for game-changing medications like HEMT. Therefore, in the present study we have prospectively assessed AS changes on a daily basis before and immediately after start of a new highly effective CFTR-modulating ETI therapy in 45 pwCF using a novel PROM, i.e. the novel CF-specific GI-symptom diary (CFAbd-day2day©). The development of the new PROM was based on the CFAbd-Score, the first CF-specific GI PROM designed and validated following FDA guidelines (17–19). To our knowledge, this is the first publication including a diary that closely captures CF-specific AS, developed following FDA guidelines and COSMIN methodology for development of a PROM (17,28).

Altogether, the results obtained with the CAbd-day2day© cumulatively calculated for each fortnight, i.e. the fortnight before ETI initiation and the second fortnight after ETI initiation, accord with our previously published results obtained with the CFAbd-Score in 107 pwCF from Germany and the UK: GI symptoms significantly decrease already after 2-4 weeks, as we previously found using the 28 item CFAbd-Score before and 4, respectively 26 weeks after initiation of ETI, retrospectively capturing the burden of GI symptoms during the preceding 14 days (22,23).

Our new findings with the CFAbd-day2day© reveal that the introduction of ETI is followed by changes in abdominal symptomatology according to quite individualized profiles, substantially differing from one pwCF to another receiving the new therapy. The extent of short-term changes in GI symptomatology is exemplified by the individual CFAbd-day2day© record from a single CF patient (Fig. 1). According to that, before introduction of the novel CFTR modulators, the patient had apparently experienced a relatively constant level in the majority of the assessed AS. Then, some symptoms like abdominal pain and bloating occurred more frequently, even from the first day of treatment with ETI. In other patients included in this study, however, some of these complaints occurred less frequently during these time frames.

Despite the highly individual pattern of changes, trends in the overall cohort are visible in Figs. 2 and 3. They reveal that some relevant symptoms appear more frequently and reveal simultaneously to have some dynamics occurring in a proportion of patients over time. For instance, abdominal pain, including its duration and intensity, increased in up to 18% of patients during the first 10 days of ETI, together with an increase in flatulence in 21% of patients on day 7. Then, after 11 to 15 days of ETI, the proportions of patients reporting pain symptoms markedly decreased to 0-10%. Also flatulence appears to improve during the last observational week (days 15-27) in many patients, together with an increase in the proportion of patients reporting a decrease of fatty and foul-smelling stool.

“Between-day variability” and “maximal absolute deviations from the median” in CFAbd-day2day© items reveal some statistically significant changes in the patterns of variability. Again, the crucial symptom of abdominal pain, including its intensity and duration, as well as “pain during bowel movements” reveal a significant reduction in the variability over the late observational period.

Notably, the present study included a relatively high proportion of younger pwCF, due to the approval of ETI in pwCF between 6 and 12 years of age carrying a F508del mutation during the study period (median age: 10 years, range: 6-55 years). According to our previous studies including the CFAbd-Score, children report significantly more often on abdominal pain, whereas adults complain significantly more often about gastroesophageal reflux (18,19). Accordingly, including a higher proportion of adult pwCF or even patients with more advanced disease progression, a different pattern of symptoms would be expected, specifically in regard to GERD and vomiting, which resulted to be rare symptoms in our cohort (16,18,19).

Calendars documenting specific patterns of symptoms are used as a golden standard in the care of patients suffering from complaints such as recurrent headache, or chronic abdominal pain, including Crohn’s disease, ulcerative colitis or irritable bowel disease. However, the pattern of AS in pwCF has been found to be rather specific due to specific CFTR deficiencies in the exocrine and endocrine pancreas, small and large intestine, as well as the bile ducts (7). Accordingly, PROMs developed for other non CF-specific abdominal pathologies like irritable bowel disease, chronic inflammatory bowel diseases, non-CF-related GER, pancreatitis, or constipation, may not be adequately sensitive for the CF-specific pattern of GI symptoms (30). In our eyes, these limitations are reflected, for instance, in the lack of sensitivity to detect ETI effects in large multicentre trials using the PAGI-SYM, PAC-SYM or PAC-QoL (27). Although significant improvements in symptoms were identified, such changes were too small to achieve clinical relevance, according to the authors of recent publications in this field. In contrast, CF-specific AS assessed with the CFAbd-Score declined in 107 pwCF from Germany and the UK from a mean of 14.9 to 10.6 pts during 24 weeks of ETI (p< 0.05), as did the five domains of pain, GERD, disorders of bowel movement, disorders of appetite and GI-related QoL (16). Likewise, a similar improvement was seen in 103 pwCF from Ireland and the UK, assessed with the CFAbd-Score during a new therapy with ETI (23). Therefore, the level of changes captured with the CF-specific CFAbd-Score can be considered as clinically relevant (10).

To our knowledge, the results presented here accord with the first study investigating AS recorded in detail after ETI initiation with a CF-specific validated diary approach. In times before HEMT approval, earlier studies assessing GI symptoms in pwCF focused on either abdominal pain with non CF-specific PROMs and/or did not report information regarding the methodology or specific content and design of implemented diaries (31–34). For instance, in 1992 Elliot et al. compared the effects of different PERT therapies on GI-complications using some kind of a symptom diary (31). They reported pwCF to prefer microcapsules as PERT because these appeared to cause lower rates of abdominal symptoms and the dosage included fewer capsules. However, no differentiation of abdominal symptoms or further information about the employed diary was provided (31). Two other studies analysed abdominal pain in pwCF with a pain diary. One investigated recurrent abdominal pain in n=8 pwCF children and the other observed the effect of nocturnal hydration on abdominal pain in n=9 pwCF (32,33). In the latter, the pain diary assessed the frequency, medications and pain intensity on a visual analogue scale, whereas the former used different PROMs for pain measurement (Eland Pain location, pain intensity measured by Faces Pain Scale-Revised (FPS-R), Mc Gill emotional status, R-CMAS anxiety score, and health-related quality of life (CF-QOL)) at the initial visit of the study (32,33). In both studies, however, further information regarding the content of the diary or its validation were missing. Recently, Van Biervliet et al. used a diary to explore effects of probiotics on GI symptoms and on the intestinal flora in n=31 pwCF. Outcomes included faecal calprotectin, pulmonary function, the nutritional status, and abdominal symptoms assessed with a diary, which queried abdominal pain, stool frequency, and treatment changes. Again, further information on the diary was not provided (34).

Diaries are especially advantageous to focus on a shorter time frame where effects or changes are expected. This applies to our setting, assessing changes of symptoms during implementation of ETI. Previously, many patients in our clinic had reported a sudden change in GI symptoms, often commencing hours after the first dosage of HEMT. Previously, the 28 items included in the CFAbd-day2day©, had been identified as highly relevant to repeatedly consulted CF patients, proxies and CF-caregivers of different professions (community voice). Overall, we saw high acceptance of our PROM.

The present results represent essential steps in the validation process of the CAbd-day2day©, revealing that the diary possesses adequate sensitivity to changes, which is defined as the ability to detect statistically significant changes after an intervention and/or treatment. Further validation steps of the CAbd-day2day© are the subject of ongoing and future studies assessing daily changes in pwCF GI symptoms in regard to relevant conditions such as CF-related diabetes (CFRD), liver involvement and DIOS among other.

In summary, implementation of the novel CFAbd-day2day© during a new therapy with the highly effective CFTR modulator ETI provides new real word insights and significant information for pwCF community and health care providers (knowledge about symptoms, expectations etc.). Furthermore, the symptom-diary can be implemented in daily care of pwCF suffering from GI symptoms to follow up their dynamics in daily life together with effects of therapeutic interventions. In addition to detecting daily changes in AS during a new HEMT, a historic game changer in CF therapy, the CFAbd-day2day© opens up further potential in CF care e.g. in following up pwCF with other specific GI symptoms. This could be essential, for instance, in pwCF suffering from DIOS, intruception, constipation, and GERD. Thereby, the subject’s pattern of symptoms, as well as effects of therapeutic interventions can be documented implementing the standardized CF-specific diary. Thereby, the CFAbd-day2day© may also help to capture critical GI complications requiring visits or transfer of pwCF to specialized CF centers. Analogous to the CFAbd-Score, the PROM is being translated to other languages and will be implemented in international studies.

## 6 Author Contributions

Conceptualization: JGM, FD. Project administration: JGM, FD, and CZ. Recruitment and Data acquisition: JGM, PH, AB, PS, LP, LB, LN, OE, SvD, PE, UGM. Analysis and interpretation of data: CZ, AB, PS, FD, and JGM. Manuscript writing: CZ, JGM, FD, PH, AB, CS, SL. All authors contributed to the article and approved the submitted version.

## Acknowledgments

We thank all individuals for their participation in this project.

## 8 Data Availability Statement

The raw data supporting the conclusions of this article will be made available by the authors, without undue reservation

## 9 Ethics

This study has been conducted in strict accordance with the ethical guidelines in the Declaration of Helsinki and, for Germany, it was approved by the Brandenburg Medical School ethics committee (Prof. Dr. Dr. Kurt J.G. Schmailzl, registration number: E-01-20200519). Furthermore, it was registered as a validation study at Clinical Trials Gov. (ClinicalTrials.gov identifier: NCT03052283)

## References

1. Ratjen F, Doring G. Cystic fibrosis. Lancet. 2003;361(9358):681–9.

2. Deutsches Mukoviszidose-Register Berichtsband 2019 [Internet]. 2020.

3. Martin C, Hamard C, Kanaan R, Boussaud V, Grenet D, Abely M, et al. Causes of death in French cystic fibrosis patients: The need for improvement in transplantation referral strategies! J Cyst Fibros. 2016;15(2):204–12.

4. Freedman SD, Wilschanski M, Schwarzenberg SJ. Advancing the GI frontier for patients with CF. Journal of cystic fibrosis: official journal of the European Cystic Fibrosis Society. 2018;17(1):1–2.

5. Ooi CY, Durie PR, Ooi CY, Durie PR. Cystic fibrosis from the gastroenterologist’s perspective. Nat Rev Gastroenterol Hepatol. 2016;13(3):175–85.

6. Munck A, Alberti C, Colombo C, Kashirskaya N, Ellemunter H, Fotoulaki M, et al. International prospective study of distal intestinal obstruction syndrome in cystic fibrosis: Associated factors and outcome. Journal of Cystic Fibrosis. 2016;15(4):531–9.

7. Ooi CY, Durie PR. Cystic fibrosis from the gastroenterologist’s perspective. Nat Rev Gastroenterol Hepatol. 2016;13(3):175–85.

8. Stefano MA, Sandy NS, Zagoya C, Duckstein F, Ribeiro AF, Mainz JG, et al. Diagnosing constipation in patients with cystic fibrosis applying ESPGHAN criteria. J Cyst Fibros. 2022;21(3):497–501.

9. Caley LR, White H, de Goffau MC, Floto RA, Parkhill J, Marsland B, et al. Cystic Fibrosis-Related Gut Dysbiosis: A Systematic Review. Dig Dis Sci. 2023.

10. Caley LR, Zagoya C, Duckstein F, White H, Shimmin D, Jones AM, et al. Diabetes is associated with increased burden of gastrointestinal symptoms in adults with cystic fibrosis. J Cyst Fibros. 2023.

11. Ramsey BW, Davies J, McElvaney NG, Tullis E, Bell SC, Drevinek P, et al. A CFTR potentiator in patients with cystic fibrosis and the G551D mutation. The New England journal of medicine. 2011;365(18):1663–72.

12. Davies JC, Wainwright CE, Canny GJ, Chilvers MA, Howenstine MS, Munck A, et al. Efficacy and safety of ivacaftor in patients aged 6 to 11 years with cystic fibrosis with a G551D mutation. Am J Respir Crit Care Med. 2013;187(11):1219–25.

13. King SJ, Tierney AC, Edgeworth D, Keating D, Williams E, Kotsimbos T, et al. Body composition and weight changes after ivacaftor treatment in adults with cystic fibrosis carrying the G551 D cystic fibrosis transmembrane conductance regulator mutation: A double-blind, placebo-controlled, randomized, crossover study with open-label extension. Nutrition. 2021;85:111124.

14. Bodewes F, Wilschanski M. CFTR Protein Function Modulation Therapy Is Finally Targeting Cystic Fibrosis-related Gastrointestinal Disease. J Pediatr Gastroenterol Nutr. 2018;66(3):372–3.

15. Davies JC, Cunningham S, Harris WT, Lapey A, Regelmann WE, Sawicki GS, et al. Safety, pharmacokinetics, and pharmacodynamics of ivacaftor in patients aged 2–5 years with cystic fibrosis and a CFTR gating mutation (KIWI): an open-label, single-arm study. The Lancet Respiratory Medicine. 2016;4(2):107–15.

16. Mainz JG, Arnold C, Hentschel J, Tabori H. Effects of Ivacaftor in Three Pediatric Siblings With Cystic Fibrosis Carrying the Mutations G551D And F508del. Arch Bronconeumol. 2018;54(4):232–4.

17. Administration USDoHaHSFaD. GUIDANCE DOCUMENT: Patient-Reported Outcome Measures: Use in Medical Product Development to Support Labeling Claims2009 10/17/2019.

18. Tabori H, Arnold C, Jaudszus A, Mentzel HJ, Renz DM, Reinsch S, et al. Abdominal symptoms in cystic fibrosis and their relation to genotype, history, clinical and laboratory findings. PLoS One. 2017;12(5):e0174463.

19. Jaudszus A, Zeman E, Jans T, Pfeifer E, Tabori H, Arnold C, et al. Validity and Reliability of a Novel Multimodal Questionnaire for the Assessment of Abdominal Symptoms in People with Cystic Fibrosis (CFAbd-Score). Patient. 2019;12(4):419–28.

20. Tabori H, Jaudszus A, Arnold C, Mentzel HJ, Lorenz M, Michl RK, et al. Relation of Ultrasound Findings and Abdominal Symptoms obtained with the CFAbd-Score in Cystic Fibrosis Patients. Sci Rep. 2017;7(1):17465.

21. Jaudszus A, Pfeifer E, Lorenz M, Beiersdorf N, Hipler UC, Zagoya C, et al. Abdominal Symptoms Assessed With the CFAbd-Score are Associated With Intestinal Inflammation in Patients With Cystic Fibrosis. J Pediatr Gastroenterol Nutr. 2022;74(3):355–60.

22. Mainz JG, Zagoya C, Polte L, Naehrlich L, Sasse L, Eickmeier O, et al. Elexacaftor-Tezacaftor-Ivacaftor Treatment Reduces Abdominal Symptoms in Cystic Fibrosis-Early results Obtained With the CF-Specific CFAbd-Score. Front Pharmacol. 2022;13:877118.

23. Mainz JG DJ, Fleming A, Elnazir B, Williamson M, McKone E, Cox D, Linnane B, Zagoya C, McNally P. Significant reduction in abdominal symptoms assessed with CFAbdscore over 4 weeks of treatment with elexacaftor/tezacaftor/ivacaftor—First results from the RECOVER study (Abstract NACFC 2021) JCF. 2021;20 Supplement 2:267.

24. Boon M, Calvo-Lerma J, Claes I, Havermans T, Asseiceira I, Bulfamante A, et al. Use of a mobile application for self-management of pancreatic enzyme replacement therapy is associated with improved gastro-intestinal related quality of life in children with Cystic Fibrosis. J Cyst Fibros. 2020;19(4):562–8.

25. Ng C, Dellschaft NS, Hoad CL, Marciani L, Ban L, Prayle AP, et al. Postprandial changes in gastrointestinal function and transit in cystic fibrosis assessed by Magnetic Resonance Imaging. J Cyst Fibros. 2021;20(4):591–7.

26. Raun AMT, Brekke G, Molgaard C, Jaudszus A, Mainz JG, Pressler T, et al. Impact of timing of PERT on gastrointestinal symptoms in Danish children and adolescents with CF. Acta Paediatr. 2022;111(2):432–9.

27. Schwarzenberg SJ, Vu PT, Skalland M, Hoffman LR, Pope C, Gelfond D, et al. Elexacaftor/tezacaftor/ivacaftor and gastrointestinal outcomes in cystic fibrosis: Report of promise-GI. J Cyst Fibros. 2022.

28. Mokkink LB, Terwee CB, Patrick DL, Alonso J, Stratford PW, Knol DL, et al. The COSMIN study reached international consensus on taxonomy, terminology, and definitions of measurement properties for health-related patient-reported outcomes. J Clin Epidemiol. 2010;63(7):737–45.

29. Bell SC, Mainz JG, MacGregor G, Madge S, Macey J, Fridman M, et al. Patient-reported outcomes in patients with cystic fibrosis with a G551D mutation on ivacaftor treatment: results from a cross-sectional study. BMC Pulm Med. 2019;19(1):146.

30. Hayee B, Watson KL, Campbell S, Simpson A, Farrell E, Hutchings P, et al. A high prevalence of chronic gastrointestinal symptoms in adults with cystic fibrosis is detected using tools already validated in other GI disorders. UNITED EUROPEAN GASTROENTEROLOGY JOURNAL. 2019;7(7):881–8.

31. Elliott RB, Escobar LC, Lees HR, Akroyd RM, Reilly HC. A comparison of two pancreatin microsphere preparations in cystic fibrosis. N Z Med J. 1992;105(930):107–8.

32. Obideen K, Wehbi M, Hoteit M, Cai Q. Nocturnal hydration--an effective modality to reduce recurrent abdominal pain and recurrent pancreatitis in patients with adult-onset cystic fibrosis. Dig Dis Sci. 2006;51(10):1744–8.

33. Munck A, Pesle A, Cunin-Roy C, Gerardin M, Ignace I, Delaisi B, et al. Recurrent abdominal pain in children with cystic fibrosis: a pilot prospective longitudinal evaluation of characteristics and management. J Cyst Fibros. 2012;11(1):46–8.

34. Van Biervliet S, Hauser B, Verhulst S, Stepman H, Delanghe J, Warzee JP, et al. Probiotics in cystic fibrosis patients: A double blind crossover placebo controlled study: Pilot study from the ESPGHAN Working Group on Pancreas/CF. Clin Nutr ESPEN. 2018;27:59–65.

